# Systematic review and critical appraisal of prediction models for diagnosis and prognosis of COVID-19 infection

**DOI:** 10.1101/2020.03.24.20041020

**Authors:** Laure Wynants, Ben Van Calster, Marc MJ Bonten, Gary S Collins, Thomas PA Debray, Maarten De Vos, Maria C. Haller, Georg Heinze, Karel GM Moons, Richard D Riley, Ewoud Schuit, Luc JM Smits, Kym IE Snell, Ewout W Steyerberg, Christine Wallisch, Maarten van Smeden

## Abstract

**Objective:** To review and critically appraise published and preprint reports of models that aim to predict either (i) presence of existing COVID-19 infection, (ii) future complications in individuals already diagnosed with COVID-19, or (iii) models to identify individuals at high risk for COVID-19 in the general population.

**Design:** Rapid systematic review and critical appraisal of prediction models for diagnosis or prognosis of COVID-19 infection.

**Data sources:** PubMed, EMBASE via Ovid, Arxiv, medRxiv and bioRxiv until 24^th^ March 2020.

**Study selection:** Studies that developed or validated a multivariable COVID-19 related prediction model. Two authors independently screened titles, abstracts and full text.

**Data extraction:** Data from included studies were extracted independently by at least two authors based on the CHARMS checklist, and risk of bias was assessed using PROBAST. Data were extracted on various domains including the participants, predictors, outcomes, data analysis, and prediction model performance.

**Results:** 2696 titles were screened. Of these, 27 studies describing 31 prediction models were included for data extraction and critical appraisal. We identified three models to predict hospital admission from pneumonia and other events (as a proxy for covid-19 pneumonia) in the general population; 18 diagnostic models to detect COVID-19 infection in symptomatic individuals (13 of which were machine learning utilising computed tomography (CT) results); and ten prognostic models for predicting mortality risk, progression to a severe state, or length of hospital stay. Only one of these studies used data on COVID-19 cases outside of China. Most reported predictors of presence of COVID-19 in suspected patients included age, body temperature, and signs and symptoms. Most reported predictors of severe prognosis in infected patients included age, sex, features derived from CT, C-reactive protein, lactic dehydrogenase, and lymphocyte count.

Estimated C-index estimates for the prediction models ranged from 0.73 to 0.81 in those for the general population (reported for all 3 general population models), from 0.81 to > 0.99 in those for diagnosis (reported for 13 of the 18 diagnostic models), and from 0.85 to 0.98 in those for prognosis (reported for 6 of the 10 prognostic models). All studies were rated at high risk of bias, mostly because of non-representative selection of control patients, exclusion of patients who had not experienced the event of interest by the end of the study, and poor statistical analysis, including high risk of model overfitting. Reporting quality varied substantially between studies. A description of the study population and intended use of the models was absent in almost all reports, and calibration of predictions was rarely assessed.

**Conclusion:** COVID-19 related prediction models are quickly entering the academic literature, to support medical decision making at a time where this is urgently needed. Our review indicates proposed models are poorly reported and at high risk of bias. Thus, their reported performance is likely optimistic and using them to support medical decision making is not advised. We call for immediate sharing of the individual participant data from COVID-19 studies to support collaborative efforts in building more rigorously developed prediction models and validating (evaluating) existing models. The aforementioned predictors identified in multiple included studies could be considered as candidate predictors for new models. We also stress the need to follow methodological guidance when developing and validating prediction models, as unreliable predictions may cause more harm than benefit when used to guide clinical decisions. Finally, studies should adhere to the TRIPOD statement to facilitate validating, appraising, advocating and clinically using the reported models.

**Systematic review registration protocol:** osf.io/ehc47/, <u>registration: osf.io/wy245</u>

**Summary boxes:** *What is already known on this topic:*

- The sharp recent increase in COVID-19 infections has put a strain on healthcare systems worldwide, necessitating efficient early detection, diagnosis of patients suspected of the infection and prognostication of COVID-19 confirmed cases.
- Viral nucleic acid testing and chest CT are standard methods for diagnosing COVID-19, but are time-consuming.
- Earlier reports suggest that the elderly, patients with comorbidity (COPD, cardiovascular disease, hypertension), and patients presenting with dyspnoea are vulnerable to more severe morbidity and mortality after COVID-19 infection.

*What this study adds:*

- We identified three models to predict hospital admission from pneumonia and other events (as a proxy for COVID-19 pneumonia) in the general population.
- We identified 18 diagnostic models for COVID-19 detection in symptomatic patients.
- 13 of these were machine learning models based on CT images.
- We identified ten prognostic models for COVID-19 infected patients, of which six aimed to predict mortality risk in confirmed or suspected COVID-19 patients, two aimed to predict progression to a severe or critical state, and two aimed to predict a hospital stay of more than 10 days from admission.
- Included studies were poorly reported compromising their subsequent appraisal, and recommendation for use in daily practice. All studies were appraised at high risk of bias, raising concern that the models may be flawed and perform poorly when applied in practice, such that their predictions may be unreliable.

## INTRODUCTION

The novel coronavirus (COVID-19) presents a significant and urgent threat to global health. Since the outbreak in early December 2019 in the Hubei Province of the People’s Republic of China, more than 775.000 cases have been confirmed in over 160 countries, and under-ascertainment of cases is likely. Over 36.000 people died from COVID-19 infection (up to 30^st^ March).^1^ Despite public health responses aimed at containing the disease and delaying the spread, several countries have been confronted with a critical care crisis, and more countries will almost certainly follow.^2-4^ Outbreaks lead to important increases in the demand for hospital beds and shortage of medical equipment, while medical staff themselves may also get infected.

To mitigate the burden on the health care system, while also providing the best possible care for patients, efficient diagnosis and prognosis is needed. Prediction models, which combine multiple predictors (variables or features) to estimate the risk of being infected or experiencing poor outcome of the infection, could assist medical staff in triaging patients when allocating limited healthcare resources. Prediction models, ranging from rule-based scoring systems to advanced machine learning models (deep learning), have already been proposed and published in response to a call to share relevant COVID-19 research findings rapidly and openly to inform the public health response and help save lives.^5^ Many of these prediction models are published in open access repositories, ahead of peer-review.

We aimed to systematically review and critically appraise currently available COVID-19 related prediction models, in particular models for diagnosis of COVID-19 in suspected cases or models for prognosis of individuals in confirmed cases. This systematic review was done in collaboration with the Cochrane Prognosis Methods group.

## METHODS

We searched PubMed, EMBASE via Ovid, bioRxiv, medRxiv, and arXiv for research on COVID-19 published after 3^rd^ January 2020. We used the publicly available publication list of the COVID-19 Living Systematic Review.^6^ This list contains studies on COVID-19 published on PubMed, EMBASE via Ovid, bioRxiv, and medRxiv, and is continuously updated. We validated the list to examine whether it is fit for purpose by comparing it to relevant hits from bioRxiv and medRxiv when combining COVID-19 search terms (covid-19, sars-cov-2, “novel corona”, 2019-ncov) with methodological search terms (diagnostic, prognostic, prediction model, machine learning, artificial intelligence, algorithm, score, deep learning, regression). All relevant hits were found on the Living Systematic Review list.^6^ We supplemented the Living Systematic Review list ^6^ with hits from PubMed searching for “covid-19”, as this was at the moment of our search not included in the Living Systematic Review ^6^ search terms for PubMed. We further supplemented the Living Systematic Review ^6^ list with studies on COVID-19 retrieved from arXiv. The search strings are listed in the Supplementary Material. In addition, we reached out to authors to include studies that were not publicly available at the time of the search, ^7 8^ and included studies that were publicly available but not on the Living Systematic Review ^6^ list at the time of our search. ^9-12^

Databases were initially searched on 13^th^ March 2020, with an update on 24^th^ March 2020. All studies were considered, regardless of language or publication status (preprint or peer reviewed articles). Studies were included if they developed or validated a multivariable model or scoring system, based on individual participant level data, to predict any COVID-19 related outcome in individuals, including to inform diagnosis, prognosis, or early identification of individuals at increased risk of developing COVID-19 pneumonia in the general population. There was no restriction on the setting (e.g., inpatients, outpatients or general population), prediction horizon (how far ahead the model predicts), included predictors, or outcomes. Epidemiological studies that aimed at modelling disease transmission or case-fatality rates, diagnostic test accuracy and predictor finding studies were excluded. Titles, abstracts and full texts were screened in duplicate for eligibility by pairs of independent reviewers (from LW, BVC, MvS), and discrepancies were resolved through discussion.

Data extraction of included articles was done by two independent reviewers (from LW, BVC, GSC, TPAD, MCH, GH, KGM, RDR, ES, LJMS, EWS, KIES, CW and MvS), using a standardized data extraction form based on the CHARMS checklist ^13^ and Prediction model Risk Of Bias ASsessment Tool (PROBAST). ^14^ We sought to extract each model’s predictive performance, using whatever measures were presented, including any summaries of discrimination (the extent to which predicted risks discriminate between participants with and without the outcome), and calibration (the extent to which predicted risks correspond to observed risks) as recommended in the TRIPOD statement. ^15^ Discrimination is often quantified by the C-index (which takes on the value of 1 in case of perfect discrimination and 0.5 is discrimination is no better than chance); calibration is often quantified by the calibration intercept (0 when the risks are not systematically over- or underestimated) and calibration slope (1 if the predicted risks are not too extreme nor too moderate). ^16^ We focus on performance statistics as estimated from the strongest available form of validation. Any discrepancies in data extraction were resolved by LW and MvS. Details on data extraction are provided in the Supplementary Material. Reporting of the article considered aspects of PRISMA ^17^ and TRIPOD ^15^.

### Patient and public involvement

It was not appropriate or possible to involve patients or the public in the design, conduct, or reporting of our research. The study protocol and preliminary results were made publicly available on osf.io/ehc47/ and medRxiv.

No ethical approval was required for the current study.

## RESULTS

A total of 2690 titles were retrieved through our systematic search (Figure 1; 1916 on 13^th^ March and an additional 774 at an update on 24^th^ March). Two additional unpublished studies were made available upon request (after a call on social media). We further included four additional studies that were publicly available but were not detected by our search. Out of 2696 titles, 85 studies were retained for abstract and full text screening. Twenty-seven studies, describing thirty-one prediction models, met the inclusion criteria and were selected for data extraction and critical appraisal. ^7-12 18-38^

**Figure 1.**
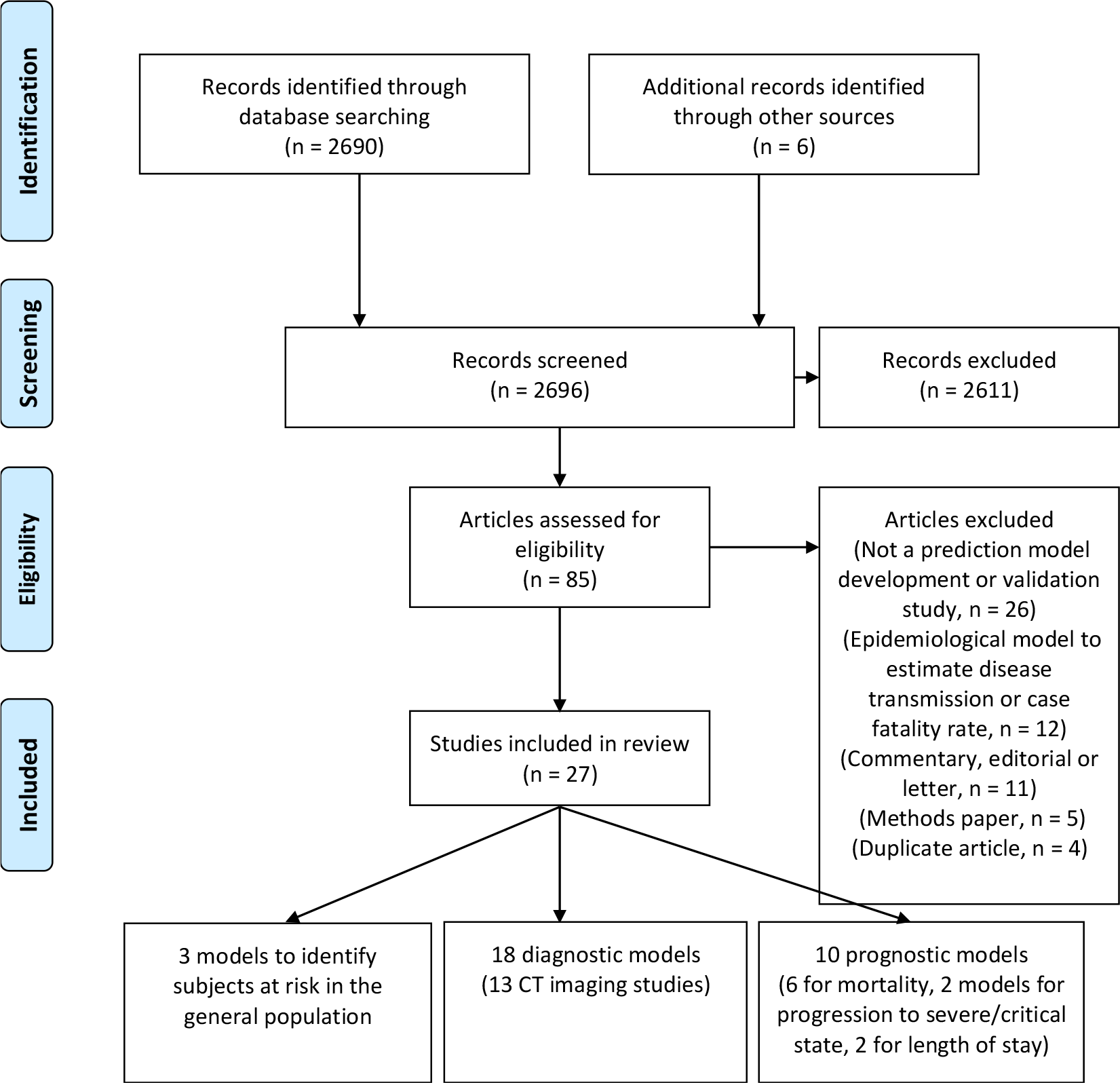
PRISMA flowchart of in- and exclusions.

### Primary datasets

Twenty-five studies used data on COVID-19 cases from China (see Supplementary Table 1), one study used data on Italian cases,^31^ and one study used international data (among others, United States, United Kingdom, China).^35^ Based on 18 of the 25 studies that reported study dates, data were collected between 8^th^ December 2019 and 15^th^ March 2020. The duration of follow-up was unclear in most studies, although one reported a median follow-up of 8.4 days,^19^ whilst another reported a median follow-up of 15 days.^37^ Some Chinese centers provided data to multiple studies, but it was unclear how much these datasets overlapped across our 25 identified studies. One study used U.S. Medicare claims data from 2015 to 2016 to estimate COVID-19 vulnerability,^8^ two studies used control CT scans from the USA or Switzerland,^11 25^ and one study used simulated data.^18^

All but one study^24^ developed prediction models for use in adults. The median age varied between studies (from 34 to 65 years, see Supplementary Table 1), as did the percentage of men (from 41% to 61%).

Among the six studies that developed prognostic models to predict mortality risk in individuals with confirmed or suspected COVID-19 infection, the percentage of deaths varied between 8% and 59% (See Table 1). This wide variation is in part due to severe sampling bias caused by studies excluding participants who still had the disease at the end of the study period (i.e., neither recovered nor died). ^7 20-22^ In addition, length of follow-up may have varied between studies (but was rarely reported), and there may be local and temporal variation in how people were diagnosed or hospitalized (and hence recruited for the studies). Among the 18 diagnostic model studies, there was only one that reported on prevalence of COVID-19 infection in those suspected of having COVID-19; the prevalence was 19% (development dataset) and 24% (validation dataset).^30^ One study reported 8% of severe cases among confirmed pediatric COVID-19 cases.^24^ Since 16 diagnostic studies used either case-control sampling or an unclear method of data collection, the prevalence in these diagnostic studies may not have been representative of their target population.

**Table 1.**
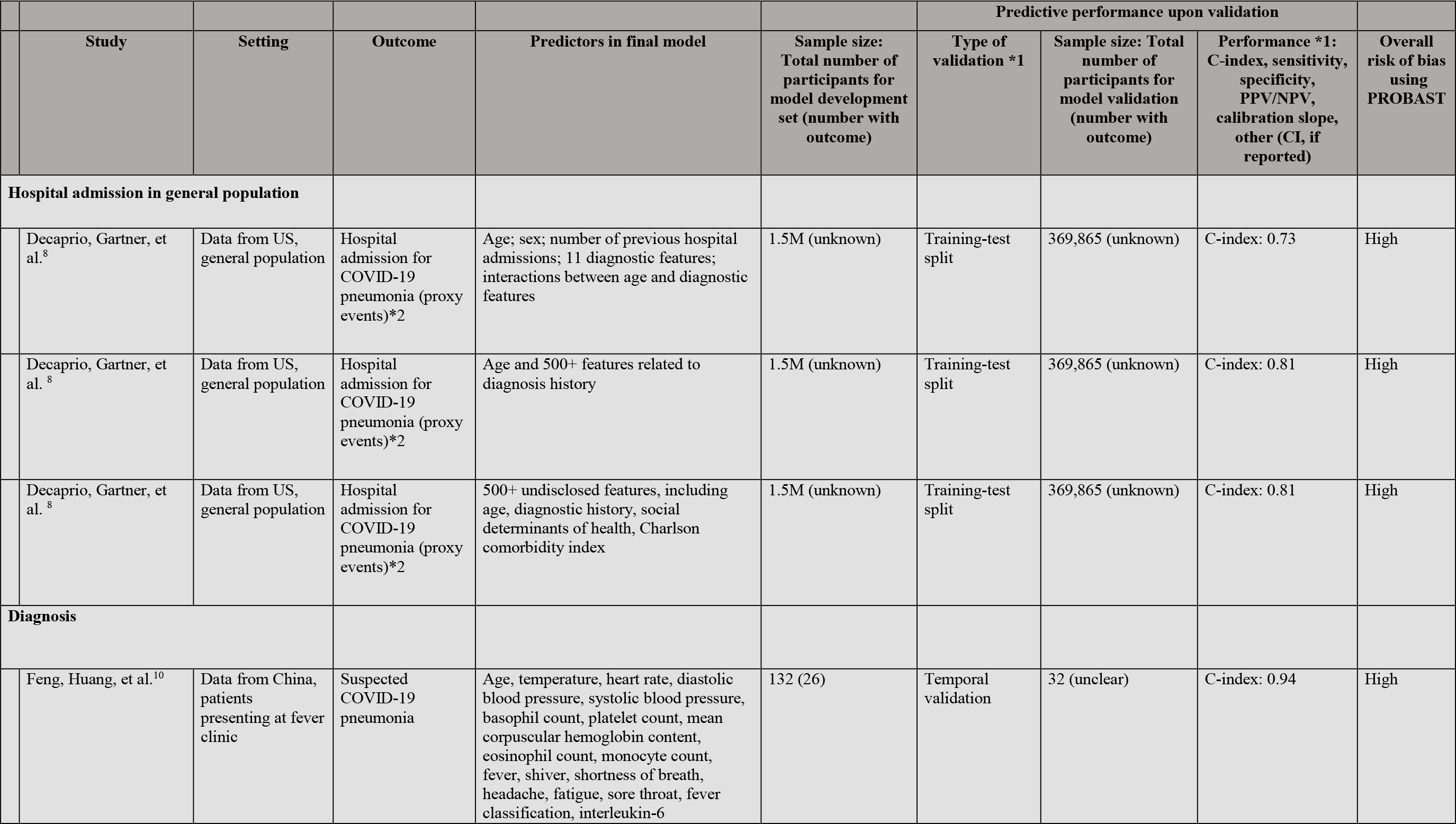
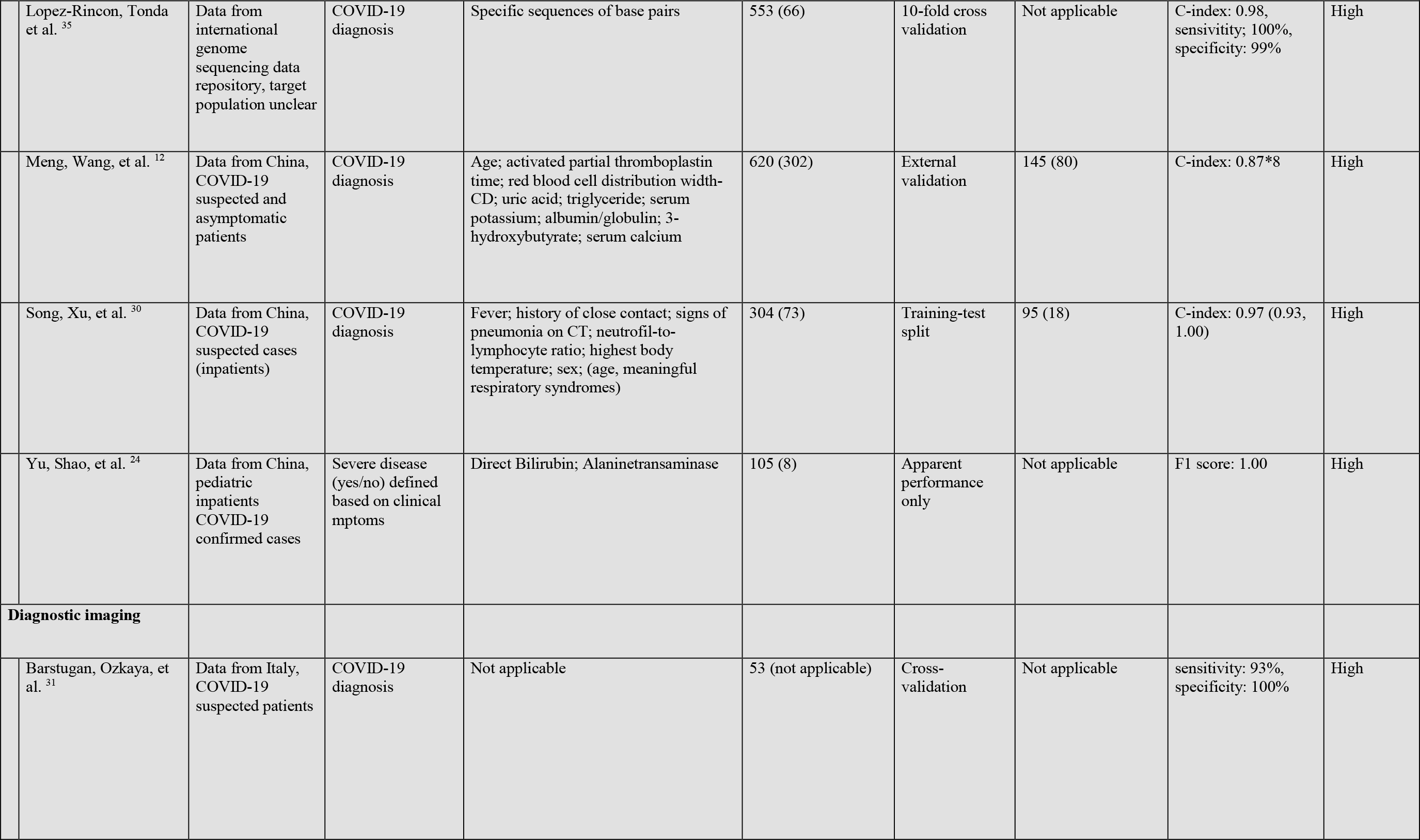
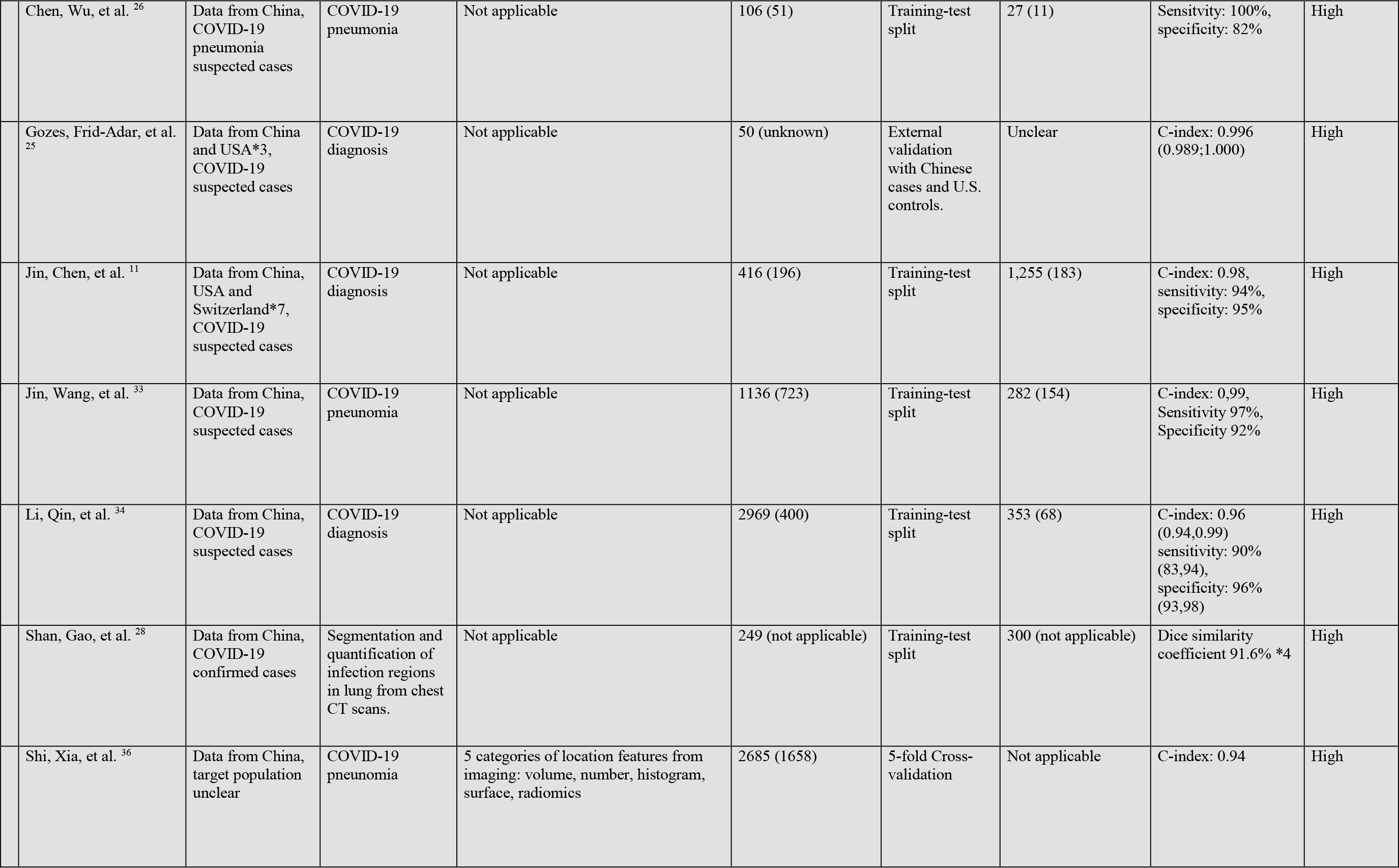
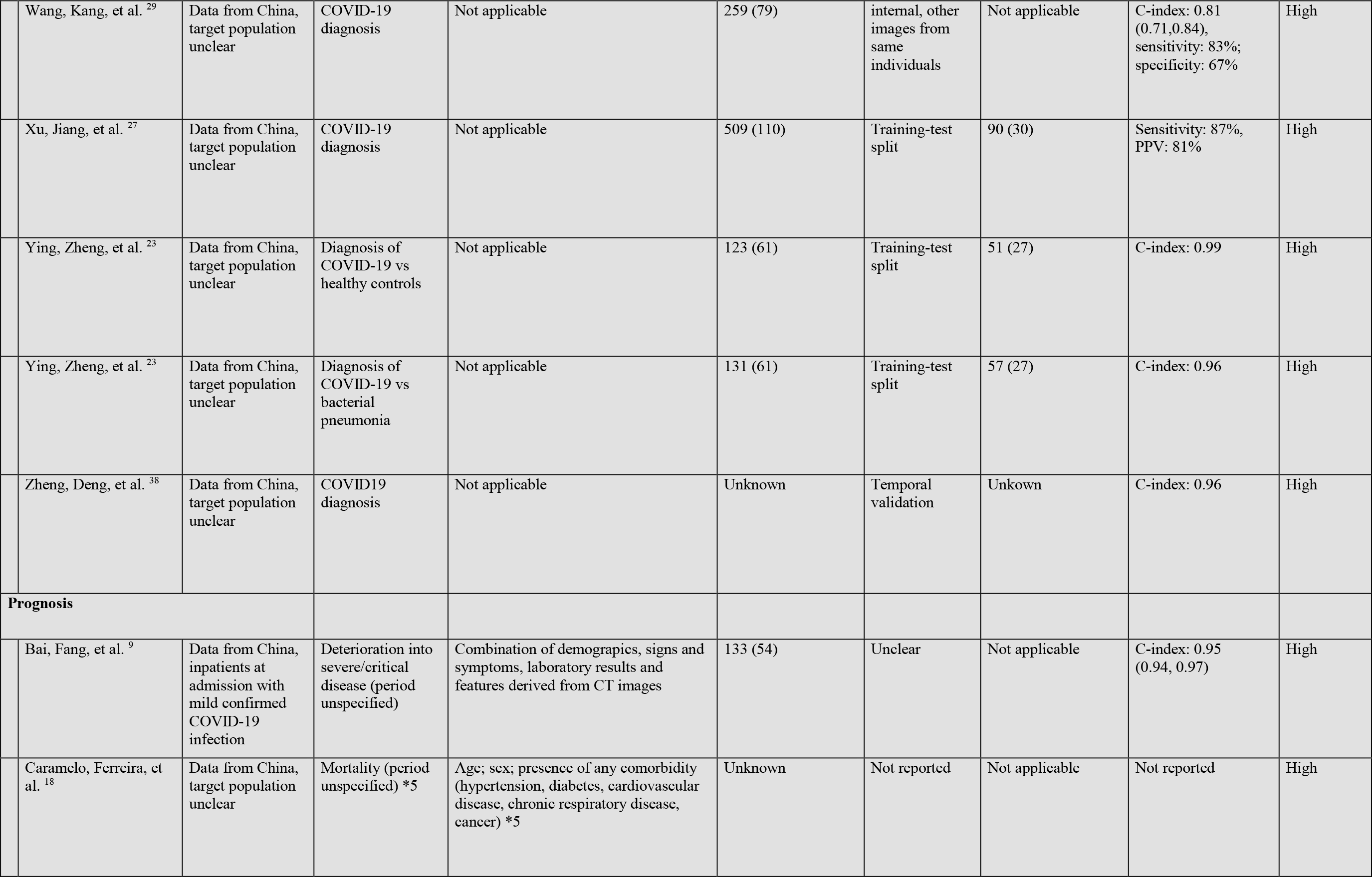
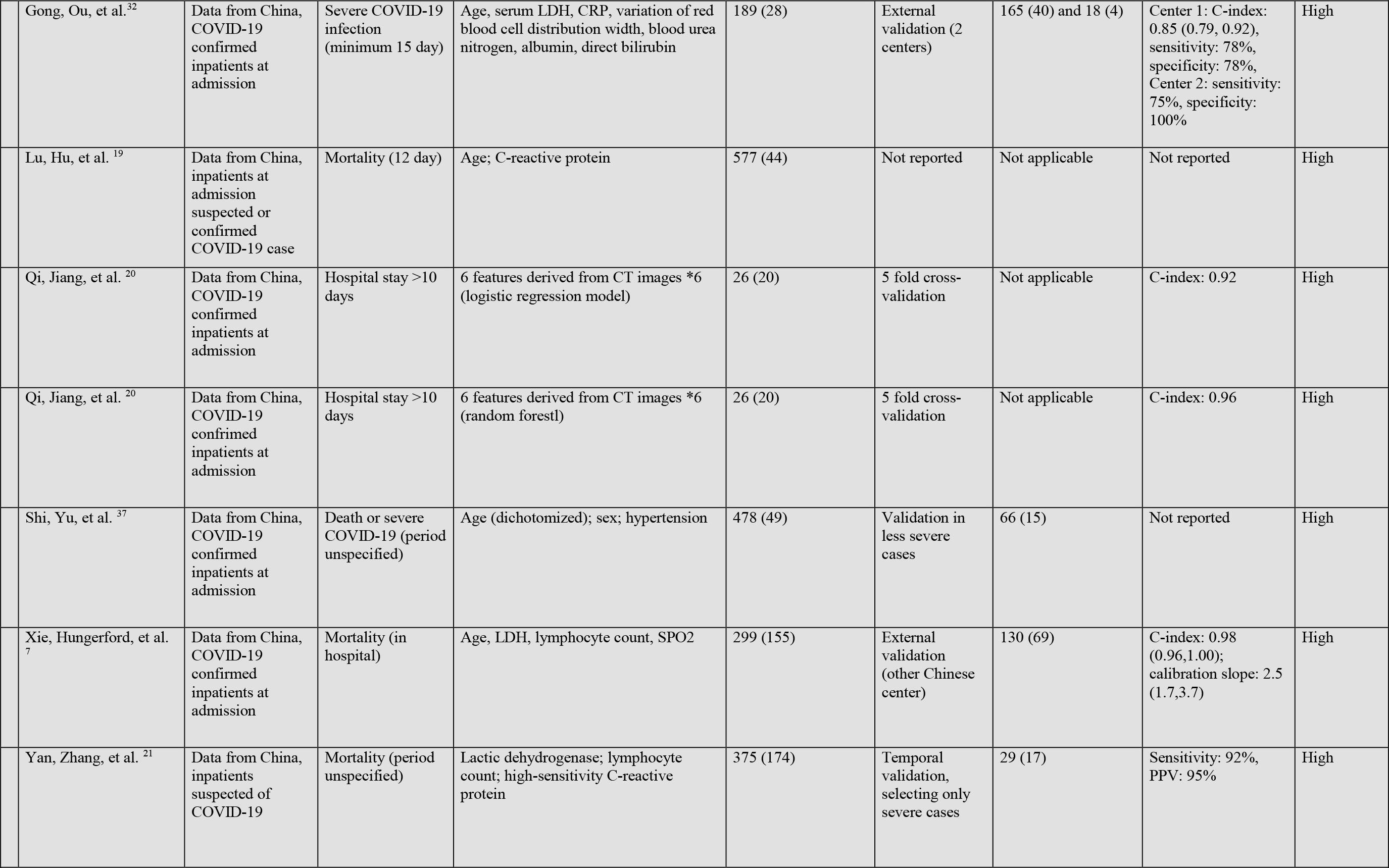
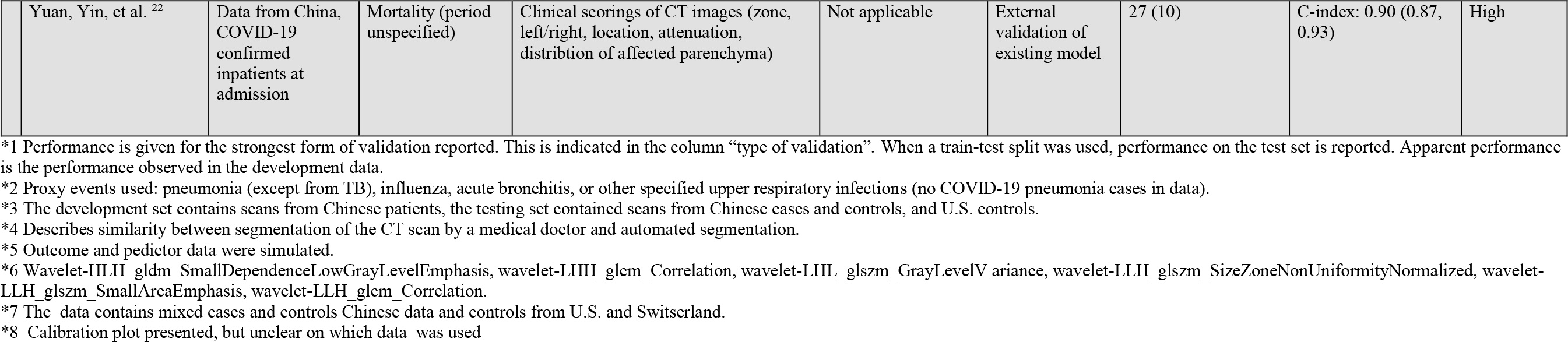
Overview of prediction models for diagnosis and prognosis of COVID-19 infection.

|  |  |  |  |  |  | Predictive performance upon validation |  |  |  |
| --- | --- | --- | --- | --- | --- | --- | --- | --- | --- |
|  | Study | Setting | Outcome | Predictors in final model | Sample size: Total number of participants for model development set (number with outcome) | Type of validation *1 | Sample size: Total number of participants for model validation (number with outcome) | Performance *1: C-index, sensitivity, specificity, PPV/NPV, calibration slope, other (CI, if reported) | Overall risk of bias using PROBAST |
| Hospital admission in general population |  |  |  |  |  |  |  |  |  |
|  | Decaprio, Gartner, et al.<sup>8</sup> | Data from US, general population | Hospital admission for COVID-19 pneumonia (proxy events)*2 | Age; sex; number of previous hospital admissions; 11 diagnostic features; interactions between age and diagnostic features | 1.5M (unknown) | Training-test split | 369,865 (unknown) | C-index: 0.73 | High |
|  | Decaprio, Gartner, et al.<sup>8</sup> | Data from US, general population | Hospital admission for COVID-19 pneumonia (proxy events)*2 | Age and 500+ features related to diagnosis history | 1.5M (unknown) | Training-test split | 369,865 (unknown) | C-index: 0.81 | High |
|  | Decaprio, Gartner, et al.<sup>8</sup> | Data from US, general population | Hospital admission for COVID-19 pneumonia (proxy events)*2 | 500+ undisclosed features, including age, diagnostic history, social determinants of health, Charlson comorbidity index | 1.5M (unknown) | Training-test split | 369,865 (unknown) | C-index: 0.81 | High |
| Diagnosis |  |  |  |  |  |  |  |  |  |
|  | Feng, Huang, et al.<sup>10</sup> | Data from China, patients presenting at fever clinic | Suspected COVID-19 pneumonia | Age, temperature, heart rate, diastolic blood pressure, systolic blood pressure, basophil count, platelet count, mean corpuscular hemoglobin content, eosinophil count, monocyte count, fever, shiver, shortness of breath, headache, fatigue, sore throat, fever classification, interleukin-6 | 132 (26) | Temporal validation | 32 (unclear) | C-index: 0.94 | High |

|  |  |  |  |  |  |  |  |  |
| --- | --- | --- | --- | --- | --- | --- | --- | --- |
| Lopez-Rincon, Tonda et al. <sup>35</sup> | Data from international genome sequencing data repository, target population unclear | COVID-19 diagnosis | Specific sequences of base pairs | 553 (66) | 10-fold cross validation | Not applicable | C-index: 0.98, sensitivity: 100%, specificity: 99% | High |
| Meng, Wang, et al. <sup>12</sup> | Data from China, COVID-19 suspected and asymptomatic patients | COVID-19 diagnosis | Age; activated partial thromboplastin time; red blood cell distribution width-CD; uric acid; triglyceride; serum potassium; albumin/globulin; 3-hydroxybutyrate; serum calcium | 620 (302) | External validation | 145 (80) | C-index: 0.87*8 | High |
| Song, Xu, et al. <sup>30</sup> | Data from China, COVID-19 suspected cases (inpatients) | COVID-19 diagnosis | Fever; history of close contact; signs of pneumonia on CT; neutrophil-to-lymphocyte ratio; highest body temperature; sex; (age, meaningful respiratory syndromes) | 304 (73) | Training-test split | 95 (18) | C-index: 0.97 (0.93, 1.00) | High |
| Yu, Shao, et al. <sup>24</sup> | Data from China, pediatric inpatients COVID-19 confirmed cases | Severe disease (yes/no) defined based on clinical mptoms | Direct Bilirubin; Alaninetransaminase | 105 (8) | Apparent performance only | Not applicable | F1 score: 1.00 | High |
| **Diagnostic imaging** |  |  |  |  |  |  |  |  |
| Barstugan, Ozkaya, et al. <sup>31</sup> | Data from Italy, COVID-19 suspected patients | COVID-19 diagnosis | Not applicable | 53 (not applicable) | Cross-validation | Not applicable | sensitivity: 93%, specificity: 100% | High |
| Chen, Wu, et al. <sup>26</sup> | Data from China, COVID-19 pneumonia suspected cases | COVID-19 pneumonia | Not applicable | 106 (51) | Training-test split | 27 (11) | Sensitivity: 100%, specificity: 82% | High |
| Gozes, Frid-Adar, et al. <sup>25</sup> | Data from China and USA*3, COVID-19 suspected cases | COVID-19 diagnosis | Not applicable | 50 (unknown) | External validation with Chinese cases and U.S. controls. | Unclear | C-index: 0.996 (0.989;1.000) | High |
| Jin, Chen, et al. <sup>11</sup> | Data from China, USA and Switzerland*7, COVID-19 suspected cases | COVID-19 diagnosis | Not applicable | 416 (196) | Training-test split | 1,255 (183) | C-index: 0.98, sensitivity: 94%, specificity: 95% | High |
| Jin, Wang, et al. <sup>33</sup> | Data from China, COVID-19 suspected cases | COVID-19 pneumonia | Not applicable | 1136 (723) | Training-test split | 282 (154) | C-index: 0.99, Sensitivity 97%, Specificity 92% | High |
| Li, Qin, et al. <sup>34</sup> | Data from China, COVID-19 suspected cases | COVID-19 diagnosis | Not applicable | 2969 (400) | Training-test split | 353 (68) | C-index: 0.96 (0.94,0.99) sensitivity: 90% (83,94), specificity: 96% (93,98) | High |
| Shan, Gao, et al. <sup>28</sup> | Data from China, COVID-19 confirmed cases | Segmentation and quantification of infection regions in lung from chest CT scans. | Not applicable | 249 (not applicable) | Training-test split | 300 (not applicable) | Dice similarity coefficient 91.6% *4 | High |
| Shi, Xia, et al. <sup>36</sup> | Data from China, target population unclear | COVID-19 pneumonia | 5 categories of location features from imaging: volume, number, histogram, surface, radiomics | 2685 (1658) | 5-fold Cross-validation | Not applicable | C-index: 0.94 | High |
|  | Wang, Kang, et al. <sup>29</sup> | Data from China, target population unclear | COVID-19 diagnosis | Not applicable | 259 (79) | internal, other images from same individuals | Not applicable | C-index: 0.81 (0.71,0.84), sensitivity: 83%; specificity: 67% | High |
|  | Xu, Jiang, et al. <sup>27</sup> | Data from China, target population unclear | COVID-19 diagnosis | Not applicable | 509 (110) | Training-test split | 90 (30) | Sensitivity: 87%, PPV: 81% | High |
|  | Ying, Zheng, et al. <sup>23</sup> | Data from China, target population unclear | Diagnosis of COVID-19 vs healthy controls | Not applicable | 123 (61) | Training-test split | 51 (27) | C-index: 0.99 | High |
|  | Ying, Zheng, et al. <sup>23</sup> | Data from China, target population unclear | Diagnosis of COVID-19 vs bacterial pneumonia | Not applicable | 131 (61) | Training-test split | 57 (27) | C-index: 0.96 | High |
|  | Zheng, Deng, et al. <sup>38</sup> | Data from China, target population unclear | COVID19 diagnosis | Not applicable | Unknown | Temporal validation | Unkown | C-index: 0.96 | High |
| **Prognosis** |  |  |  |  |  |  |  |  |  |
|  | Bai, Fang, et al. <sup>9</sup> | Data from China, inpatients at admission with mild confirmed COVID-19 infection | Deterioration into severe/critical disease (period unspecified) | Combination of demographics, signs and symptoms, laboratory results and features derived from CT images | 133 (54) | Unclear | Not applicable | C-index: 0.95 (0.94, 0.97) | High |
|  | Caramelo, Ferreira, et al. <sup>18</sup> | Data from China, target population unclear | Mortality (period unspecified) *5 | Age; sex; presence of any comorbidity (hypertension, diabetes, cardiovascular disease, chronic respiratory disease, cancer) *5 | Unknown | Not reported | Not applicable | Not reported | High |

|  |  |  |  |  |  |  |  |  |
| --- | --- | --- | --- | --- | --- | --- | --- | --- |
| Gong, Ou, et al.<sup>32</sup> | Data from China, COVID-19 confirmed inpatients at admission | Severe COVID-19 infection (minimum 15 day) | Age, serum LDH, CRP, variation of red blood cell distribution width, blood urea nitrogen, albumin, direct bilirubin | 189 (28) | External validation (2 centers) | 165 (40) and 18 (4) | Center 1: C-index: 0.85 (0.79, 0.92), sensitivity: 78%, specificity: 78%, Center 2: sensitivity: 75%, specificity: 100% | High |
| Lu, Hu, et al.<sup>19</sup> | Data from China, inpatients at admission suspected or confirmed COVID-19 case | Mortality (12 day) | Age; C-reactive protein | 577 (44) | Not reported | Not applicable | Not reported | High |
| Qi, Jiang, et al.<sup>20</sup> | Data from China, COVID-19 confirmed inpatients at admission | Hospital stay >10 days | 6 features derived from CT images *6 (logistic regression model) | 26 (20) | 5 fold cross-validation | Not applicable | C-index: 0.92 | High |
| Qi, Jiang, et al.<sup>20</sup> | Data from China, COVID-19 confirmed inpatients at admission | Hospital stay >10 days | 6 features derived from CT images *6 (random forest) | 26 (20) | 5 fold cross-validation | Not applicable | C-index: 0.96 | High |
| Shi, Yu, et al.<sup>37</sup> | Data from China, COVID-19 confirmed inpatients at admission | Death or severe COVID-19 (period unspecified) | Age (dichotomized); sex; hypertension | 478 (49) | Validation in less severe cases | 66 (15) | Not reported | High |
| Xie, Hungerford, et al.<sup>7</sup> | Data from China, COVID-19 confirmed inpatients at admission | Mortality (in hospital) | Age, LDH, lymphocyte count, SPO2 | 299 (155) | External validation (other Chinese center) | 130 (69) | C-index: 0.98 (0.96,1.00); calibration slope: 2.5 (1.7,3.7) | High |
| Yan, Zhang, et al.<sup>21</sup> | Data from China, inpatients suspected of COVID-19 | Mortality (period unspecified) | Lactic dehydrogenase; lymphocyte count; high-sensitivity C-reactive protein | 375 (174) | Temporal validation, selecting only severe cases | 29 (17) | Sensitivity: 92%, PPV: 95% | High |
|  | Yuan, Yin, et al. <sup>22</sup> | Data from China, COVID-19 confirmed inpatients at admission | Mortality (period unspecified) | Clinical scorings of CT images (zone, left/right, location, attenuation, distribution of affected parenchyma) | Not applicable | External validation of existing model | 27 (10) | C-index: 0.90 (0.87, 0.93) | High |

\*1 Performance is given for the strongest form of validation reported. This is indicated in the column “type of validation”. When a train-test split was used, performance on the test set is reported. Apparent performance is the performance observed in the development data.
\*2 Proxy events used: pneumonia (except from TB), influenza, acute bronchitis, or other specified upper respiratory infections (no COVID-19 pneumonia cases in data).
\*3 The development set contains scans from Chinese patients, the testing set contained scans from Chinese cases and controls, and U.S. controls.
\*4 Describes similarity between segmentation of the CT scan by a medical doctor and automated segmentation.
\*5 Outcome and predictor data were simulated.
\*6 Wavelet-HLH\_gldm\_SmallDependenceLowGrayLevelEmphasis, wavelet-LHH\_glcmm\_Correlation, wavelet-LHL\_glszm\_GrayLevelVariance, wavelet-LLH\_glszm\_SizeZoneNonUniformityNormalized, wavelet-LLH\_glszm\_SmallAreaEmphasis, wavelet-LLH\_glcmm\_Correlation.
\*7 The data contains mixed cases and controls Chinese data and controls from U.S. and Switzerland.
\*8 Calibration plot presented, but unclear on which data was used

In what follows, we give an overview of the 31 prediction models reported in the 27 identified studies (Table 1). Modeling details are provided in Supplementary Table 2, and the availability of models in a format for use in clinical practice is discussed in Box 1.

#### Box 1. Availability of the models in a format for use in clinical practice

Twelve studies presented their models in a format for use in clinical practice. However, because all models were at high risk of bias, we do not recommend their routine use before they are properly externally validated.

##### Models to predict hospital admission for COVID-19 pneumonia in the general Population

The “CV-19 vulnerability index” to detect hospital admission for COVID-19 pneumonia from other respiratory infections (e.g. pneumonia, influenza), is available as an online tool.^8 67^

##### Diagnostic models

The “COVID-19 Diagnosis Aid APP” is available on iOS and android devices to diagnose suspected and asymptomatic patients.^12^ The “suspected COVID-19 pneumonia Diagnosis Aid System” is available as an online tool.^10 68^ The “COVID-19 Early Warning Score” to detect COVID-19 infection in adults is available as a score chart in an article.^30^ A decision tree to detect severe disease for pediatric COVID-19 confirmed patients is also available in an article.^24^

##### Diagnostic models based on CT imaging

Three of the seven AI models to assist with diagnosis based on CT results, are available via web applications. ^23 26 29 69-71^ One model is deployed in 16 hospitals, but the authors do not provide any usable tools in their study.^33^

##### Prognostic models

To assist in the prognosis of mortality, a nomogram (a graphic aid to calculate mortality risk),^7^ a decision tree,^21^ and a CT-based scoring rule are available in the articles.^22^There is also a nomogram to predict progression to severe COVID-19.^32^ Five studies made their source code available on GitHub.^8 11 34 35 38^ Ten studies did not include any usable equation, format or reference for use or validation of their prediction model.

### Models to predict the risk of hospital admission due to COVID-19 pneumonia in the general population

Three models predicted the risk of hospital admission for COVID-19 pneumonia for individuals in the general population, but used admission due to non-tuberculosis pneumonia, influenza, acute bronchitis, or upper respiratory infections as outcomes in a dataset without any COVID-19 cases (see Table 1).^8^ Among the predictors were age, sex, previous hospital admissions, comorbidity data, and social determinants of health. The study estimated C-indices of 0.73, 0.81 and 0.81 for the three models.

### Diagnostic models to detect COVID-19 infection in symptomatic individuals

One study developed a model to detect COVID-19 pneumonia in fever clinic patients (estimated C-index 0.94),^10^ one to diagnose COVID-19 in suspected cases (estimated C-index 0.97),^30^ one to diagnose COVID-19 in suspected and asymptomatic cases (estimated C-index 0.87),^12^ one to diagnose COVID-19 using deep learning of genomic sequences (estimated C-index 0.98),^35^ and one to diagnose severe disease in symptomatic paediatric inpatients based on direct bilirubin and alaninetransaminase (reporting an F1 score of 1.00, indicating 100% observed sensitivity and specificity).^24^ Only one study reported assessing calibration, but it was unclear how this was done.^12^ Predictors used in more than one model were age (n=3), body temperature or fever (n=2), and signs and symptoms (such as shortness of breath, headache, shiver, sore throat, fatigue) (n=2) (see Table 1).

Thirteen prediction models were proposed to support the diagnosis of COVID-19 or COVID-19 pneumonia (and monitor progression) based on CT images. The predictive performance varied widely, with estimated C-index values ranging from 0.81 to nearly 1.

### Prognostic models for patients diagnosed with COVID-19 infection

We identified ten prognostic models (Table 1). Of these, six estimated mortality risk in suspected or confirmed COVID-19 patients.^7 18 19 21 22 37^ The intended use of these models (namely when to use it, in whom to use it, and the prediction horizon, e.g., mortality by what time) was not clearly described. Two models aimed to predict a hospital stay of more than 10 days from admission.^20^ Two models aimed to predict progression to a severe or critical state.^9 32^ Predictors included in more than one prognostic model were age (n=5), sex (n=2), features derived from CT-scoring (n=5), C-reactive protein (n=3), lactic dehydrogenase (n=3), and lymphocyte count (n=2) (see Table 1).

Only two studies predicting mortality reported a C-index; they obtained estimates of 0.90 ^22^ and 0.98 ^7^. One study also evaluated calibration.^7^ When applied to new patients, their model yielded probabilities of mortality that were too high for low-risk patients and too low for high-risk patients (calibration slope >1), despite excellent discrimination.^7^ One study developed two models to predict a hospital stay of >10 days and estimated C-indices of 0.92 and 0.96.^20^ The two studies that developed models to predict progression to a severe or critical state estimated C-indices of 0.95 and 0.85.^9 32^ One of these also reported perfect calibration, but it was unclear how this was evaluated. ^32^

### Risk of bias

All models were at high risk of bias according to assessment with PROBAST (Table 1), which suggests that their predictive performance when used in practice is likely lower than what is reported, and so gives concern that their predictions are unreliable. Details on common causes for risk of bias are given in Box 2 for each type of model.

#### Box 2. Common causes of risk of bias in the 19 reported prediction models.

##### Models to predict hospital admission for COVID-19 pneumonia in the general Population

These models were based on Medicare claims data, and used proxy outcomes to predict hospital admission for COVID-19 pneumonia, in absence of COVID-19 cases.^8^

##### Diagnostic models

Individuals without COVID-19 (or a portion thereof) were excluded, altering the disease prevalence.^30^ Controls had viral pneumonia, which is not representative of the target population for a screening model.^12^ The test used to determine the outcome varied between participants,^12^ or one of the predictors (fever) was part of the outcome definition.^10^ Predictors were dichotomized, leading to a loss of information.^24 30 36^

##### Diagnostic models based on CT imaging

There was generally poor reporting on which patients’ CT images were obtained during clinical routine, and it was unclear whether the selection of controls was sampled from the target population (i.e., patients suspected of COVID-19).^11 23 29 33 36^ It was often unclear how regions of interest (ROIs) were annotated. Images were sometimes annotated by only one scorer without quality control ^25 27^, the model output influenced annotation^28^, or the “ground truth” which was used to build the model was a composite outcome based on the same CT images used to make the prediction, among other things.^38^ Careful description of model specification and subsequent estimation was lacking, challenging the transparency and reproducibility of the models. Every study used a different deep learning architecture, including established and specifically designed ones, without benchmarking the used architecture with respect to others.

##### Prognostic models

Study participants were often simply excluded because they did not develop the outcome at the end of the study period but were still in follow-up (i.e., in the hospital and neither recovered nor died), yielding a highly selected study sample.^7 20-22^ In addition, only one study accounted for censoring by using Cox regression.^19^ One study developed a model to predict future severity using cross-sectional data (i.e., the participants already were severely ill at inclusion).^37^ This implies that the timing of the measurement of the predictors is not appropriate, and the (unclearly defined) outcome may have been influenced by the predictor values. Other studies used highly subjective predictors,^22^ or the last available predictor measurement from electronic health records (rather than the measurement of the predictor value at the time the model is intended to be used).^21^ Dichotomization of predictors was often applied which tends to lead to loss of information.^24 30^

Eleven of the twenty-seven studies had a high risk of bias for the “participants” domain (Table 2), indicating that the participants enrolled in the studies may not be representative for the models’ targeted populations. Unclear reporting on the inclusion of participants prohibited a risk of bias assessment in eight studies. Four out of twenty-seven studies had a high risk of bias for the “predictors” domain, indicating that predictors were not available at the models’ intended time of use, not clearly defined, or influenced by the outcome measurement. The diagnostic model studies that used CT imaging predictors were all scored as “unclear” on the “predictors” domain. The publications often lacked clear information on the preprocessing steps (e.g., cropping of images). Moreover, complex machine learning algorithms transform CT images into predictors in an intransparent way, which makes it challenging to fully apply the PROBAST predictors section for such imaging studies. Most studies used outcomes that are easy to assess (e.g., death, presence of COVID-19 by laboratory confirmation). Nonetheless, there was reason to be concerned of bias induced by the outcome measurement in ten studies, due to the use of subjective or proxy-outcomes (e.g., non COVID-19 severe respiratory infections).

**Table 2.** Risk of bias assessment (using PROBAST) based on four domains across 27 studies creating prediction models for COVID-19

| Authors | Risk of bias: participants | Risk of bias: predictors | Risk of bias: outcome | Risk of bias: analysis |
| --- | --- | --- | --- | --- |
| **Hospital admission in general population** |  |  |  |  |
| DeCaprio, Gartner, et al. <sup>8</sup> | high | low | high | high |
| **Diagnosis** |  |  |  |  |
| Feng, Huang, et al. <sup>10</sup> | low | unclear | high | high |
| Lopez-Rincon, Tonda, et al. <sup>35</sup> | unclear | low | low | high |
| Meng, Wang, et al. <sup>12</sup> | high | low | high | high |
| Song, Xu, et al. <sup>30</sup> | high | unclear | low | high |
| Yu, Shao, et al. <sup>24</sup> | unclear | unclear | unclear | high |
| **Diagnostic imaging** |  |  |  |  |
| Barstugan, Ozkaya, et al. <sup>31</sup> | unclear | unclear | unclear | high |
| Chen, Wu, et al. <sup>26</sup> | high | unclear | low | high <sup>*1</sup> |
| Gozes, Frid-Adar, et al. <sup>25</sup> | unclear | unclear | high | high |
| Jin, Chen, et al. <sup>11</sup> | high | unclear | unclear | high <sup>*2</sup> |
| Jin, Wang, et al. <sup>33</sup> | high | unclear | high | high <sup>*1</sup> |
| Li, Qin, et al. <sup>34</sup> | low | unclear | low | high |
| Shan, Gao, et al. <sup>28</sup> | unclear | unclear | high | high <sup>*2</sup> |
| Shi, Xia, et al. <sup>36</sup> | high | unclear | low | high |
| Wang, Kang, et al. <sup>29</sup> | high | unclear | low | high |
| Xu, Jiang, et al. <sup>27</sup> | high | unclear | high | high |
| Ying, Zheng, et al. <sup>23</sup> | unclear | unclear | low | high |
| Zheng, Deng, et al. <sup>38</sup> | unclear | unclear | high | high |
| **Prognosis** |  |  |  |  |
| Bai, Fang, et al. <sup>9</sup> | low | unclear | unclear | high |
| Caramelo, Ferreira, et al. <sup>18</sup> | high | high | high | high |
| Gong, Ou, et al. <sup>32</sup> | low | unclear | unclear | high |
| Lu, Hu, et al. <sup>19</sup> | low | low | low | high |
| Qi, Jiang, et al. <sup>20</sup> | unclear | low | low | high |
| Shi, Yu, et al. <sup>37</sup> | high | high | high | high |
| Xie, Hungerford, et al. <sup>7</sup> | low | low | low | high |
| Yan, Zhang, et al. <sup>21</sup> | low | high | low | high |
| Yuan, Yin, et al. <sup>22</sup> | low | high | low | high |

<sup>\*1</sup> Risk of bias high due to not evaluating calibration. If this criterion is not taken into account, analysis risk of bias would have been unclear.
<sup>\*2</sup> Risk of bias high due to not evaluating calibration. If this criterion is not taken into account, analysis risk of bias would have been low.

All studies were at high risk of bias for the “analysis” domain (Table 2). Many studies had small sample sizes (Table 1), leading to an increased risk of overfitting, particularly if complex modeling strategies were used. Three studies did not report the predictive performance of the developed model, and one study reported only the apparent performance (that is, the performance in the exact same data as was used to develop the model, without adjustment for optimism due to potential overfitting).

Four models were externally validated in the model development study (i.e., in an independent dataset, excluding random train-test splits and temporal splits).^7 12 25 32^ However, in three of these studies, the external validation datasets are likely unrepresentative of the target population (Box 2).^7 12 25^ Consequently, predictive performance may be different if the model were applied in the target population. Gong, Ou, et al had a satisfactory predictive performance on two unbiased but small external validation datasets.^32^ One study was a small (n=27) external validation reporting satisfactory predictive performance of a model originally developed for avian influenza H7N9 pneumonia, but patients that had not recovered at the end of the study period were excluded, leading to a selection bias.^22^ Only three studies assessed calibration, ^7 12 32^ but this was likely done suboptimally in two studies.^12 32^

## DISCUSSION

### Main findings

In this systematic review of prediction models related to the COVID-19 pandemic, we identified and critically appraised 27 studies that described 31 prediction models for detecting individuals at risk for hospital admission for COVID-19 pneumonia in the general population, for diagnosis of COVID-19 in symptomatic individuals, and for prognosis of COVID-19 infected patients. All models reported good to even excellent predictive performance, but all were appraised as high risk of bias, due to a combination of poor reporting and poor methodological conduct for participant selection, predictor description and statistical methods used. As expected, in these early COVID-19 related prediction model studies, clinical data from COVID-19 patients is still scarce and limited to data from China, Italy, and international registries. With few exceptions, the available sample size and number of events for the outcomes of interest were limited, which is a known problem for building prediction models, increasing the risk of overfitting the prediction model.^39^ A high risk of bias implies that these models are likely to perform worse in practice than the performance that is reported by the researchers. Hence, the estimated C-indices, often close to 1 and indicating near-perfect discrimination, are highly likely to be optimistic. Five studies carried out an external validation, ^7 12 25 32 22^ and only one study assessed calibration correctly.^7^

We reviewed thirteen studies that used advanced machine learning methodology on chest CT scans to diagnose COVID-19 disease, COVID-19 related pneumonia, or to assist in segmentation of lung images. The predictive performance measures showed a high to almost perfect ability to identify COVID-19, although these models and their evaluations also suffered from a high risk of bias, notably due to poor reporting and an artificial mix of COVID-19 cases and non-cases.

### Challenges and opportunities

The main aim of prediction models is to support medical decision making. It is therefore key to identify a target population in which predictions serve a clinical need, and a representative dataset (preferably comprising consecutive patients) on which the prediction model can be developed and validated. This target population must also be carefully described such that the performance of the developed or validated model can be appraised in context, and users know in which individuals the model can be applied to make predictions. However, the included studies in our systematic review often lacked an adequate description of the study population, which leaves users of these models in doubt of the models’ applicability. While we recognize that all studies were done under severe time constraints caused by urgency, we recommend that any studies currently in preprint and all future studies should adhere to the TRIPOD reporting guideline^15^ to improve the description of their study population as well as their modeling choices. TRIPOD translations (e.g., in Chinese and Japanese) are also available at www.tripod-statement.org.

A better description of the study population may also help understand the observed variability in the reported outcomes across studies, such as COVID-19 related mortality. The variability in the relative frequencies of the predicted outcomes presents an important challenge to the prediction modeler: a prediction model applied in a setting with a different relative frequency of the outcome may produce predictions that are miscalibrated ^40^ and may need to be updated before it can safely be applied in that new setting.^16 41^ Indeed, such an update may often be required when prediction models are transported to different healthcare systems, which requires COVID-19 patient data to be available from that system.

COVID-19 prediction problems will often not present as a simple binary classification task. Complexities in the data should be handled appropriately. For example, a prediction horizon should be specified for prognostic outcomes (e.g., 30-day mortality). If study participants have neither recovered nor died within that time period, their data should not be excluded from analysis, as most reviewed studies have done. Instead, an appropriate time-to-event analysis should be considered to allow for administrative censoring.^16^ It should be noted that censoring due to other reasons, for instance due to quick recovery and loss to follow-up of patients that are no longer at risk of death from COVID-19, may necessitate analysis in a competing risk framework.^42^

Instead of developing and updating predictions in their local setting, Individual Participant Data (IPD) from multiple countries and healthcare systems may facilitate better understanding of the generalizability and implementation prediction models across different settings and populations, and may greatly improve their applicability and robustness in routine care.^43-47^

The evidence base for the development and validation of prediction models related to COVID-19 will quickly increase over the coming months. Together with the increasing evidence from predictor finding studies ^48-54^ and open peer review initiatives for COVID-19 related publications,^55^ data registries ^56-60^ are being set up. To maximize the new opportunities and to facilitate IPD meta-analyses, the WHO has recently released a new data platform to encourage sharing of anonymized COVID-19 clinical data.^61^ To leverage the full potential of these evolutions, international and interdisciplinary collaboration in terms of data acquisition and model building is crucial.

### Limitations of this study

With new publications on COVID-19 related prediction models that are currently quickly entering the medical literature, this systematic review cannot be viewed as an up-to-date list of all currently available COVID-19 related prediction models. Also, 24 of the studies we reviewed were only available as a preprint, and they might improve after peer review, when entering the official medical literature. We have also found other prediction models which are currently implemented in clinical practice without scientific publications ^62^ and web risk calculators launched for use while the scientific manuscript was still under review (and unavailable upon request).^63^ These unpublished models naturally fall outside the scope of this review of the literature.

### Implications for practice

All 31 reviewed prediction models were found to have a high risk of bias and evidence from independent external validation of these models is currently lacking. However, the urgency of diagnostic and prognostic models to assist in quick and efficient triage of patients in the COVID-19 pandemic may encourage clinicians to implement prediction models without sufficient documentation and validation. Although we cannot let perfect be the enemy of good, earlier studies have shown that models were of limited use in the context of a pandemic,^64^ and they may even cause more harm than good.^65^ Hence, we cannot recommend any model for use in practice at this point.

We anticipate that more COVID-19 data on the individual participant level will soon become available. These could be used to validate and update currently available prediction models.^16^ For example, one model predicting progression to severe COVID-19 within 15 days after admission showed promising discrimination when validated externally on two small but unselected cohorts.^32^ As reporting in this study was insufficiently detailed and the validation was in small Chinese datasets, validation in larger, international datasets is needed. Due to differences between health care systems (e.g., Chinese and European) in admission, discharge, and testing criteria for patients with COVID-19, we anticipate most existing models will need to be updated (i.e., adjusted to the local setting).

When building a new prediction model, it is recommended to build on previous literature and expert opinion to select predictors, rather than selecting predictors in a purely data-driven way ^16^. This is especially true for datasets with limited sample size.^66^ Based on the predictors included in multiple models identified by our review, we encourage researchers to consider incorporating the following as candidate predictors: (i) for diagnostic models - age, body temperature, and (respiratory) signs and symptoms; (ii) for prognostic models - age, sex, C-reactive protein, lactic dehydrogenase, lymphocyte count, and potentially features derived from CT-scoring. Predictors that were included in both a diagnostic and a prognostic model were albumin (or albumin/globin), direct bilirubin, and red blood cell distribution width; these could be considered as well. By pointing to the most important methodological challenges and issues in design and reporting of the currently available models, we hope to have provided a useful starting point for further studies aiming at developing new models or validating and updating existing ones.

This systematic review aims to be the first stage of a living review of this field, in collaboration with the Cochrane Prognosis Methods Group. We will update this review and appraisal continuously, to provide up-to date information for healthcare decision makers and professionals, as more international research emerges over time.

## CONCLUSION

Diagnostic and prognostic models for COVID-19 are available and they all appear to show good to excellent discriminative performance. However, these models are at high risk of bias mainly due to non-representative selection of control patients, exclusion of patients who had not experienced the event of interest by the end of the study, and model overfitting. Hence, their performance estimates are likely to be optimistic and misleading. Future studies should address these concerns. Sharing data and expertise for development, validation and updating of COVID-19 related prediction models is urgently needed.

## Data Availability

Extracted data is partially reported in Tables 1-2 and Supplementary Tables 1-2. The original manuscripts are published open access (links provided in the reference list).
The protocol for this systematic review was registered on OSF and publicly available(osf.io/ehc47/).

## ACKNOWLEDGMENTS

We thank the authors who made their work available, by posting it on public registries or sharing it confidentially.

## FOOTNOTES

### Contributors

LW conceived the study; LW and MvS designed the study; LW, MvS and BVC screened titles and abstracts for inclusion. LW, BVC, GSC, TPAD, MCH, GH, KGM, RDR, ES, LJMS, EWS, KIES, CW and MvS extracted and analysed data. MD helped interpret the findings on deep learning studies and MB and MCH assisted in the interpretation from a clinical viewpoint. LW and MvS wrote the first draft, which all authors revised for critical content. All authors approved the final manuscript. LW and MvS are the guarantors. The corresponding author attests that all listed authors meet authorship criteria and that no others meeting the criteria have been omitted.

## REFERENCES

1. Dong E, Du H, Gardner L. An interactive web-based dashboard to track COVID-19 in real time. Lancet Infect Dis 2020 doi: 10.1016/s1473-3099(20)30120-1

2. Arabi YM, Murthy S, Webb S. COVID-19: a novel coronavirus and a novel challenge for critical care. Intensive Care Med 2020 doi: 10.1007/s00134-020-05955-1

3. Grasselli G, Pesenti A, Cecconi M. Critical Care Utilization for the COVID-19 Outbreak in Lombardy, Italy: Early Experience and Forecast During an Emergency Response. JAMA 2020 doi: 10.1001/jama.2020.4031

4. Xie J, Tong Z, Guan X, et al. Critical care crisis and some recommendations during the COVID-19 epidemic in China. Intensive Care Med 2020 doi: 10.1007/s00134-020-05979-7

5. Sharing research data and findings relevant to the novel coronavirus (COVID-19) outbreak [Available from: https://wellcome.ac.uk/press-release/sharing-research-data-and-findings-relevant-novel-coronavirus-covid-19-outbreak accessed 22.03.2020.]

6. Living Evidence on COVID-19 [Available from: https://ispmbern.github.io/covid-9/living-review/index.html accessed 22.03.2020.]

7. Xie J, Hungerford D, Chen H, et al. Development and external validation of a prognostic multivariable model on admission for hospitalized patients with COVID-19. medRxiv 2020:2020.03.28.20045997. doi: 10.1101/2020.03.28.20045997

8. DeCaprio D, Gartner J, Burgess T, et al. Building a COVID-19 Vulnerability Index. arXiv e-prints 2020. https://ui.adsabs.harvard.edu/abs/2020arXiv200307347D.

9. Bai X, Fang C, Zhou Y, et al. Predicting COVID-19 malignant progression with AI techniques. medRxiv 2020:2020.03.20.20037325. doi: 10.1101/2020.03.20.20037325

10. Feng C, Huang Z, Wang L, et al. A Novel Triage Tool of Artificial Intelligence Assisted Diagnosis Aid System for Suspected COVID-19 pneumonia In Fever Clinics. medRxiv 2020:2020.03.19.20039099. doi: 10.1101/2020.03.19.20039099

11. Jin C, Chen W, Cao Y, et al. Development and Evaluation of an AI System for COVID-19 Diagnosis. medRxiv 2020:2020.03.20.20039834. doi: 10.1101/2020.03.20.20039834

12. Meng Z, Wang M, Song H, et al. Development and utilization of an intelligent application for aiding COVID-19 diagnosis. medRxiv 2020:2020.03.18.20035816. doi: 10.1101/2020.03.18.20035816

13. Moons KG, de Groot JA, Bouwmeester W, et al. Critical appraisal and data extraction for systematic reviews of prediction modelling studies: the CHARMS checklist. PLoS Med 2014;11(10):e1001744. doi: 10.1371/journal.pmed.1001744

14. Moons KGM, Wolff RF, Riley RD, et al. PROBAST: A Tool to Assess Risk of Bias and Applicability of Prediction Model Studies: Explanation and Elaboration. Ann Intern Med 2019;170(1):W1–w33. doi: 10.7326/m18-1377

15. Moons KGM, Altman DG, Reitsma JB, et al. Transparent Reporting of a multivariable prediction model for Individual Prognosis Or Diagnosis (TRIPOD): Explanation and ElaborationThe TRIPOD Statement: Explanation and Elaboration. Ann Intern Med 2015;162(1):W1–W73. doi: 10.7326/M14-0698

16. Steyerberg EW. Clinical Prediction Models: A Practical Approach to Development, Validation, and Updating. New York, NY: Springer US 2019.

17. Liberati A, Altman DG, Tetzlaff J, et al. The PRISMA statement for reporting systematic reviews and meta-analyses of studies that evaluate healthcare interventions: explanation and elaboration. PLoS Med 2009;6 doi: 10.1371/journal.pmed.1000100

18. Caramelo F, Ferreira N, Oliveiros B. Estimation of risk factors for COVID-19 mortality - preliminary results. medRxiv 2020:2020.02.24.20027268. doi: 10.1101/2020.02.24.20027268

19. Lu J, Hu S, Fan R, et al. ACP risk grade: a simple mortality index for patients with confirmed or suspected severe acute respiratory syndrome coronavirus 2 disease (COVID-19) during the early stage of outbreak in Wuhan, China. medRxiv 2020:2020.02.20.20025510. doi: 10.1101/2020.02.20.20025510

20. Qi X, Jiang Z, Yu Q, et al. Machine learning-based CT radiomics model for predicting hospital stay in patients with pneumonia associated with SARS-CoV-2 infection: A multicenter study. medRxiv 2020:2020.02.29.20029603. doi: 10.1101/2020.02.29.20029603

21. Yan L, Zhang H-T, Xiao Y, et al. Prediction of criticality in patients with severe Covid-19 infection using three clinical features: a machine learning-based prognostic model with clinical data in Wuhan. medRxiv 2020:2020.02.27.20028027. doi: 10.1101/2020.02.27.20028027

22. Yuan M, Yin W, Tao Z, et al. Association of radiologic findings with mortality of patients infected with 2019 novel coronavirus in Wuhan, China. PLoS One 2020;15(3):e0230548. doi: 10.1371/journal.pone.0230548

23. Song Y, Zheng S, Li L, et al. Deep learning Enables Accurate Diagnosis of Novel Coronavirus (COVID-19) with CT images. medRxiv 2020:2020.02.23.20026930. doi: 10.1101/2020.02.23.20026930

24. Yu H, Shao J, Guo Y, et al. Data-driven discovery of clinical routes for severity detection in COVID-19 pediatric cases. medRxiv 2020:2020.03.09.20032219. doi: 10.1101/2020.03.09.20032219

25. Gozes O, Frid-Adar M, Greenspan H, et al. Rapid AI Development Cycle for the Coronavirus (COVID-19) Pandemic: Initial Results for Automated Detection & Patient Monitoring using Deep Learning CT Image Analysis. arXiv e-prints 2020. https://ui.adsabs.harvard.edu/abs/2020arXiv200305037G

26. Chen J, Wu L, Zhang J, et al. Deep learning-based model for detecting 2019 novel coronavirus pneumonia on high-resolution computed tomography: a prospective study. medRxiv 2020:2020.02.25.20021568. doi: 10.1101/2020.02.25.20021568

27. Xu X, Jiang X, Ma C, et al. Deep Learning System to Screen Coronavirus Disease 2019 Pneumonia. arXiv e-prints 2020. https://ui.adsabs.harvard.edu/abs/2020arXiv200209334X

28. Shan F, Gao Y, Wang J, et al. Lung Infection Quantification of COVID-19 in CT Images with Deep Learning. arXiv e-prints 2020. https://ui.adsabs.harvard.edu/abs/2020arXiv200304655S

29. Wang S, Kang B, Ma J, et al. A deep learning algorithm using CT images to screen for Corona Virus Disease (COVID-19). medRxiv 2020:2020.02.14.20023028. doi: 10.1101/2020.02.14.20023028

30. Song C-Y, Xu J, He J-Q, et al. COVID-19 early warning score: a multi-parameter screening tool to identify highly suspected patients. medRxiv 2020:2020.03.05.20031906. doi: 10.1101/2020.03.05.20031906

31. Barstugan M, Ozkaya U, Ozturk S. Coronavirus (COVID-19) Classification using CT Images by Machine Learning Methods. arXiv e-prints 2020. https://ui.adsabs.harvard.edu/abs/2020arXiv200309424B

32. Gong J, Ou J, Qiu X, et al. A Tool to Early Predict Severe 2019-Novel Coronavirus Pneumonia (COVID-19) : A Multicenter Study using the Risk Nomogram in Wuhan and Guangdong, China. medRxiv 2020:2020.03.17.20037515. doi: 10.1101/2020.03.17.20037515

33. Jin S, Wang B, Xu H, et al. AI-assisted CT imaging analysis for COVID-19 screening: Building and deploying a medical AI system in four weeks. medRxiv 2020:2020.03.19.20039354. doi: 10.1101/2020.03.19.20039354

34. Li L, Qin L, Xu Z, et al. Artificial Intelligence Distinguishes COVID-19 from Community Acquired Pneumonia on Chest CT. Radiology 2020:200905. doi: 10.1148/radiol.2020200905

35. Lopez-Rincon A, Tonda A, Mendoza-Maldonado L, et al. Accurate Identification of SARS-CoV-2 from Viral Genome Sequences using Deep Learning. bioRxiv 2020:2020.03.13.990242. doi: 10.1101/2020.03.13.990242

36. Shi F, Xia L, Shan F, et al. Large-Scale Screening of COVID-19 from Community Acquired Pneumonia using Infection Size-Aware Classification. arXiv e-prints 2020. https://ui.adsabs.harvard.edu/abs/2020arXiv200309860S

37. Shi Y, Yu X, Zhao H, et al. Host susceptibility to severe COVID-19 and establishment of a host risk score: findings of 487 cases outside Wuhan. Crit Care 2020;24(1):108. doi: 10.1186/s13054-020-2833-7

38. Zheng C, Deng X, Fu Q, et al. Deep Learning-based Detection for COVID-19 from Chest CT using Weak Label. medRxiv 2020:2020.03.12.20027185. doi: 10.1101/2020.03.12.20027185

39. Riley RD, Ensor J, Snell KIE, et al. Calculating the sample size required for developing a clinical prediction model. BMJ 2020;368:m441. doi: 10.1136/bmj.m441

40. Van Calster B, McLernon DJ, van Smeden M, et al. Calibration: the Achilles heel of predictive analytics. BMC Med 2019;17(1):1–7.

41. Steyerberg EW. Clinical Prediction Models: A Practical Approach to Development, Validation, and Updating. New York, NY: Springer US 2009.

42. Austin PC, Lee DS, Fine JP. Introduction to the Analysis of Survival Data in the Presence of Competing Risks. Circulation 2016;133(6):601–9. doi: 10.1161/circulationaha.115.017719

43. Riley RD, Ensor J, Snell KI, et al. External validation of clinical prediction models using big datasets from e-health records or IPD meta-analysis: opportunities and challenges. BMJ 2016;353:i3140. doi: 10.1136/bmj.i3140

44. Debray TP, Riley RD, Rovers MM, et al. Individual participant data (IPD) meta-analyses of diagnostic and prognostic modeling studies: guidance on their use. PLoS Med 2015;12(10):e1001886. doi: 10.1371/journal.pmed.1001886

45. Steyerberg EW, Harrell FE, Jr. Prediction models need appropriate internal, internal-external, and external validation. J Clin Epidemiol 2016;69:245–7. doi: 10.1016/j.jclinepi.2015.04.005

46. Wynants L, Kent DM, Timmerman D, et al. Untapped potential of multicenter studies: a review of cardiovascular risk prediction models revealed inappropriate analyses and wide variation in reporting. Diagn Progn Res 2019;3:6. doi: 10.1186/s41512-019-0046-9

47. Wynants L, Riley RD, Timmerman D, et al. Random-effects meta-analysis of the clinical utility of tests and prediction models. Stat Med 2018;37(12):2034–52. doi: 10.1002/sim.7653

48. Zhou F, Yu T, Du R, et al. Clinical course and risk factors for mortality of adult inpatients with COVID-19 in Wuhan, China: a retrospective cohort study. Lancet 2020 doi: 10.1016/s0140-6736(20)30566-3

49. Li K, Wu J, Wu F, et al. The Clinical and Chest CT Features Associated with Severe and Critical COVID-19 Pneumonia. Invest Radiol 2020 doi: 10.1097/rli.0000000000000672

50. Li B, Yang J, Zhao F, et al. Prevalence and impact of cardiovascular metabolic diseases on COVID-19 in China. Clin Res Cardiol 2020 doi: 10.1007/s00392-020-01626-9

51. Jain V, Yuan J-M. Systematic review and meta-analysis of predictive symptoms and comorbidities for severe COVID-19 infection. medRxiv 2020:2020.03.15.20035360. doi: 10.1101/2020.03.15.20035360

52. Rodriguez-Morales AJ, Cardona-Ospina JA, Gutierrez-Ocampo E, et al. Clinical, laboratory and imaging features of COVID-19: A systematic review and meta-analysis. Travel Med Infect Dis 2020:101623. doi: 10.1016/j.tmaid.2020.101623

53. Lippi G, Plebani M, Henry BM. Thrombocytopenia is associated with severe coronavirus disease 2019 (COVID-19) infections: A meta-analysis. Clin Chim Acta 2020;506:145–48. doi: https://doi.org/10.1016/j.cca.2020.03.022

54. Zhao X, Zhang B, Li P, et al. Incidence, clinical characteristics and prognostic factor of patients with COVID-19: a systematic review and meta-analysis. medRxiv 2020:2020.03.17.20037572. doi: 10.1101/2020.03.17.20037572

55. Johansson M, Saderi D. Open peer-review platform for COVID-19 preprints. Nature 2020;579(7797):29.

56. Xu B, Kraemer MU, Gutierrez B, et al. Open access epidemiological data from the COVID-19 outbreak. The Lancet Infectious Diseases 2020. doi: 10.1016/s1473-3099(20)30119-5

57. COVID-19 DATABASE [Available from: https://www.sirm.org/category/senza-categoria/covid-19/ accessed 22.03.2020.]

58. Kaggle Datasets [Available from: https://www.kaggle.com/datasets accessed 22.03.2020.]

59. Covid chestxray dataset [Available from: https://github.com/ieee8023/covid-chestxray-dataset accessed 22.03.2020.

60. European registry of patients with COVID-19 including cardiovascular risk and complications [Available from: https://capacity-covid.eu/ accessed 28.03.2020.]

61. Coronavirus disease (COVID-19) technical guidance: Early investigations protocols [Available from: https://www.who.int/emergencies/diseases/novel-coronavirus-2019/technical-guidance/early-investigations accessed 22.03.2020.]

62. Infervision [Available from: https://global.infervision.com/ accessed 22.03.2020.]

63. COVID-19 Response Center [Available from: <https://surgisphere.com/covid-19-response-center/> accessed 22.03.2020.]

64. Enfield K, Miller R, Rice T, et al. Limited Utility of SOFA and APACHE II Prediction Models for ICU Triage in Pandemic Influenza. Chest 2011;140(4):913A. doi: 10.1378/chest.1118087

65. Van Calster B, Vickers AJ. Calibration of risk prediction models: impact on decision-analytic performance. Med Decis Making 2015;35(2):162–9. doi: 10.1177/0272989x14547233

66. van Smeden M, Moons KG, de Groot JA, et al. Sample size for binary logistic prediction models: Beyond events per variable criteria. Stat Methods Med Res 2018:962280218784726. doi: 10.1177/0962280218784726

67. The COVID-19 Vulnerability Index (CV19 Index) [Available from: https://closedloop.ai/cv19index/ accessed 22.03.2020.]

68. Suspected COVID-19 pneumonia Diagnosis Aid System [Available from: https://intensivecare.shinyapps.io/COVID19/. accessed 28.03.2020.]

69. AI diagnostic system for 2019-nCoV [Available from: http://121.40.75.149/znyx-ncov/index accessed 22.03.2020.]

70. ai.nscc [Available from: https://ai.nscc-tj.cn/thai/deploy/public/pneumonia_ct accessed 22.03.2020.]

71. Discriminating COVID-19 Pneumonia from CT Images [Available from: http://biomed.nscc-gz.cn/server/Ncov2019 accessed 22.03.2020.

